## Supplementary figures and images for "3D finite element models reveal regional fatty infiltration modulates tibialis anterior force generating capacity in FSHD"

### 3D plots of regional fat infiltration patterns

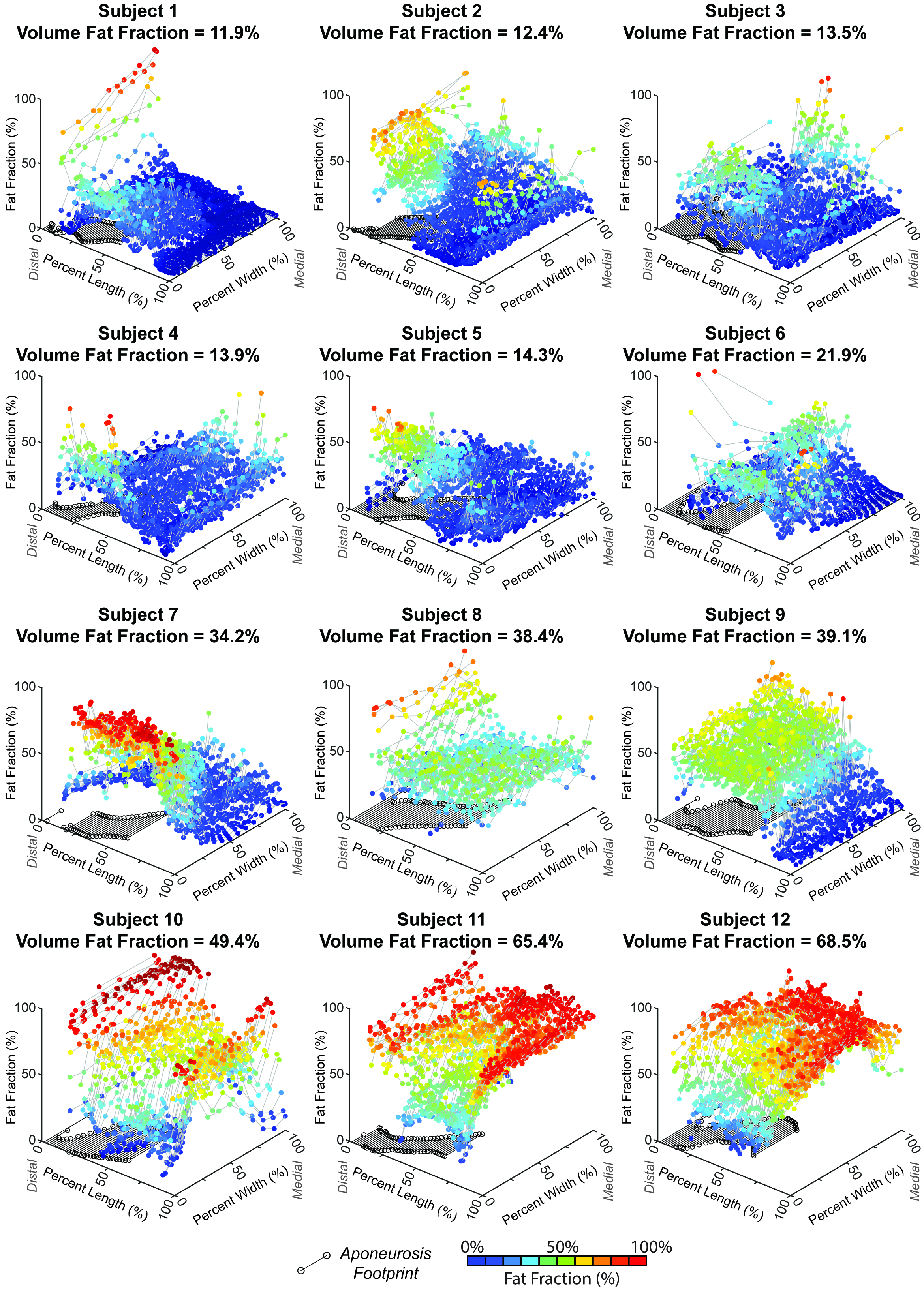

### CSA Fat Fraction along Length of Muscle

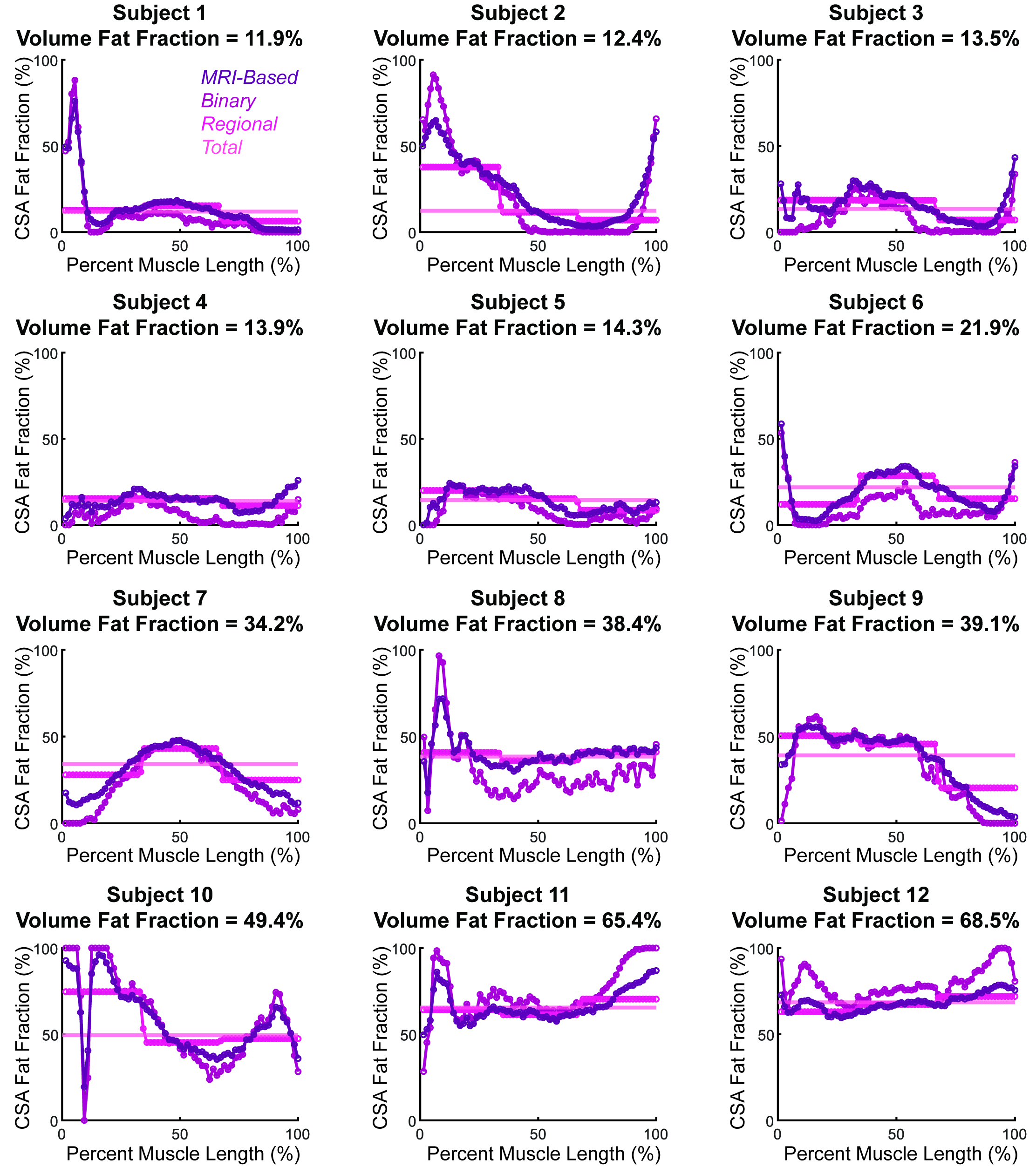
